# Direct evidence of alteration to the human neural intracellular trafficking system following exposure to HSV: A systematic review

**DOI:** 10.1101/2025.01.09.25320268

**Authors:** Michael B. Vaughan, Dante J. Bellai, Mark G. Rae

## Abstract

There is a growing interest in the potential role that lifelong chronic viral infections may play in the aetiology of certain neurodegenerative diseases. For example, the disruption of neuronal intracellular trafficking by repeated reactivations of latent herpes viruses has been proposed as a possible inductive agent of Alzheimer’s disease (AD). This systematic review examines the extent of primary data that have examined the effect(s) of herpes viruses on the human intraneuronal trafficking (HINT) system. Three databases (MEDLINE, SCOPUS, and Web of Science), were repeatedly searched. 5782 articles were identified, of which 13 met our eligibility criteria, and five were included for qualitative synthesis. Four of the five collated studies presented data from cell culture experiments, wherein herpes simplex virus 1 (HSV-1) altered lysosomal load, lysosomal function, and activity-regulated cytoskeleton-associated protein (ARC) mRNA/protein expression in a cell-type/species specific manner. One of the five studies was a clinical case study that presented observations on a fatal herpes encephalopathy, wherein the neuronal endoplasmic reticulum and Golgi apparatus were ablated by the rampant infection. Unfortunately, there is an extremely limited amount of primary research examining the effects of herpes viruses on the HINT system, but within that limited pool there is clear evidence of cell type-/stage- specific responses to infection. Future studies investigating herpes infection in neurodegenerative disease need to utilise cell-type and species- specific models if they are to have any hope of extrapolating to a clinical setting.

## Introduction

### Rationale

Human herpes viruses are a collection of DNA viruses that are defined by their abil- ity to remain dormant within host cells between periods of symptomatic infection [1, 2].

Herpes simplex viruses 1 and 2 (HSV-1, HSV-2) are estimated to infect 66.6% (0-49 years of age) and 13.2% (15-49 years of age) of the global population respectively [3].

Most commonly associated with the oral or genital lesions that occur during periods of active infections, both HSV subtypes can survive within sensory neurons for the lifetime of infected individuals [4]. Following primary infection of the oral or genital mucosa, vi- ral replicants undergo retrograde microtubule-associated transport along axons of the sensory ganglia innervating infected tissue [5, 6]. These neurons then either undergo pro- ductive lytic infection, establish a latent infection, or achieve some intermediate state be- tween the two [7]. Lytic infection involves a well characterised process of viral replication- associated gene expression, while viral latency is a process of near total viral gene repres- sion [8, 9]. In recent years there is growing recognition that viral reactivation, observed *via* the detection of viral shedding, occurs more commonly than mucosal symptom pre- sentation [10, 11]. Similarly, it is now appreciated that rather than the viral genome remaining “silent” within neurons during periods of latency, there are in fact repeated attempted reactivations that are quelled before active viral replication commences [12, 13]. Such continuous activity begs the question; what effect does continual pathogen transcription, replication and suppression have on neurons?

While herpes simplex encephalitis is a recognised rare acute infection of the central ner- vous system (1 in 250000 - 500000 cases per year) that is primarily characterised by in-flammation [14, 15], the pathology of chronic neurotropic infection with HSV is not well understood. The possible involvement of differing herpes viruses in the development of Alzheimer’s disease (AD) is tentatively supported by recent findings, such as the presence of HSV in amyloid plaques [16], the apparent neuroprotective effects of herpesviridae antivirals in reducing risk of dementia development [17, 18], and associations between HSV-1 infection susceptibility and the AD risk factor allele APOE-*ɛ*4 [19–21]. Multiple re- views and talks are available outlining the full evidence for and against this Herpes-AD hypothesis [22–24].

Within our lab, the role of intracellular trafficking disruption in the aetiology of certain neurodegenerative conditions is of particular interest. Several well-known neurodegen- erative diseases are caused by protein misfolding or genetic mutations affecting com- ponents of the vesicle trafficking system, such as microtubule integrity and endosome mobility [25](Fig.1 A). For example, AD is characterised by the accumulation and aggre- gation of amyloid *β* (A*β*) and hyperphosphorylated tau proteins, Parkinson’s disease (PD) is associated with the misfolding of alpha-synuclein, and Huntington’s disease is caused by a genetic mutation leading to the expansion of CAG repeats in the HTT gene, resulting in the production of a misfolded huntingtin protein; Each misfolded protein disrupts intra- cellular trafficking, leading to neuronal cell death [26–28]. Conversely, amyotrophic lat- eral sclerosis, Charcot-Marie-Tooth Disease, and hereditary spastic paraplegia, are each caused by mutations in genes encoding trafficking components (TDP-43, RAB7A, and SPG4 respectively) [29–31]. This is notable as the herpes viruses utilise the intracellular trafficking system for viral entry and egress [32](Fig.1b). This usage of intracellular com- ponents, paired with near ubiquitous population seropositivity and the lifelong nature of the infections, has led to the belief that repeated disruption of the human intraneuronal trafficking (HINT) system by HSV over several decades could instigate neurodegenerative processes.

**Figure 1.**
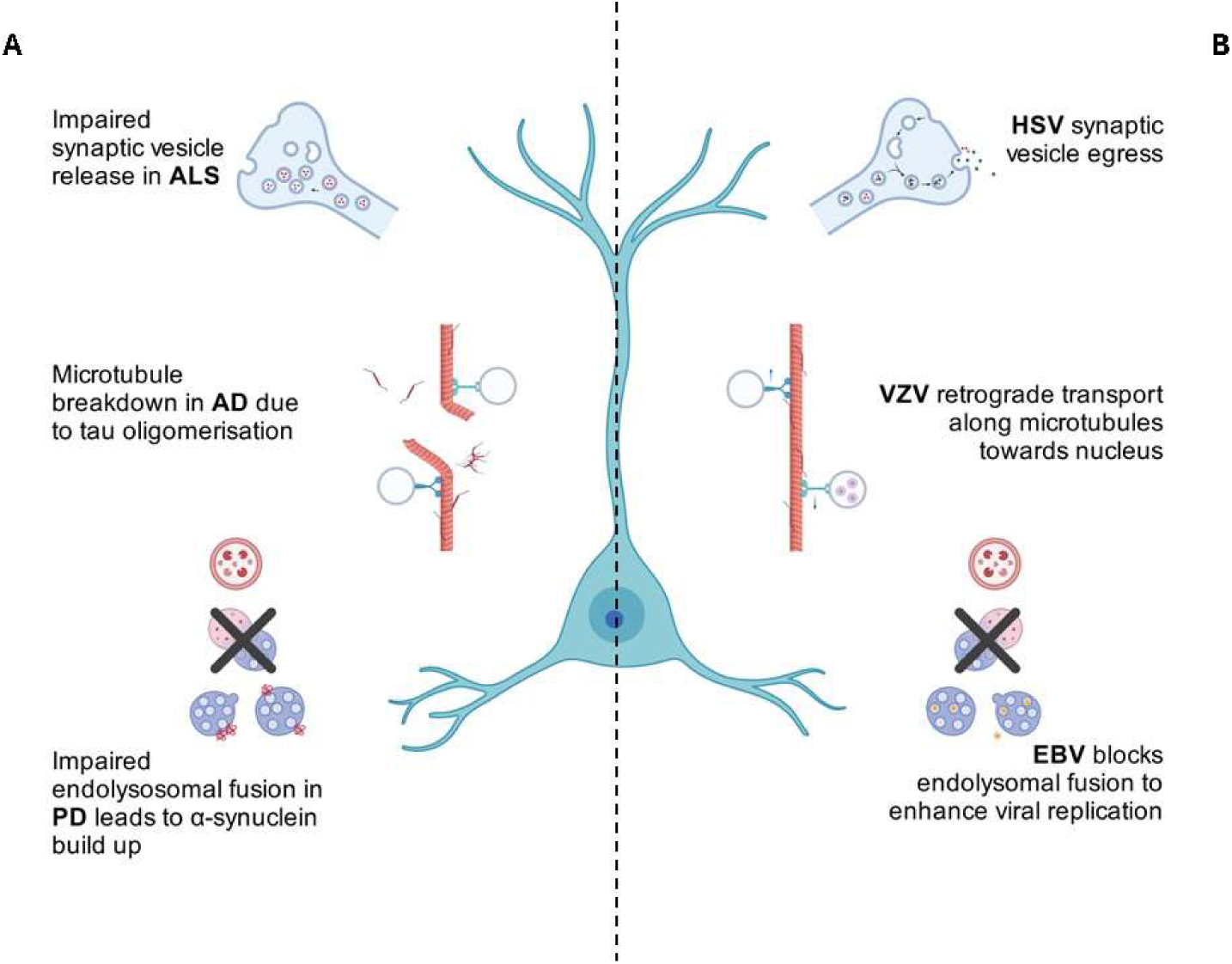
**HINT & Herpes** *Commonly recognized intracellular trafficking elements that are disrupted by neurodegeneration (A) and herpes viruses infection (B)*.

### Objectives

The primary objective of this systematic review of the literature was to investigate the known/confirmed effects of HSV on the HINT system. The secondary objective was to identify specific alterations in HINT components induced by HSV for future studies into the possible relation between viral infection and neurodegenerative diseases.

Given the human-specific nature of neurodegenerative diseases and the limitations of aetiological discovery in animal studies to date, we have focused exclusively on primary research assessing the intracellular trafficking system during and following HSV infection in human neuronal cells.

### Research Question

Is there any evidence of alterations to the neural endosomal network of humans after exposure to HSV-1 and/or HSV-2 when compared to controls (unaffected cells)?

## Methods

The search strategy was jointly designed by MBV and DJB and agreed upon with MGR. The search, selection of studies, and data extraction were performed independently by both MBV and DJB. Agreement on final report inclusion was achieved after review of the full-text articles that met eligibility criteria by both MBV and DJB. Final data synthesis and generation of the current manuscript was carried out by MBV and MGR in line with PRISMA 2020 guidelines[33].

### Search Strategy

The initial search strategy design for this study used the population, intervention, com- parison and outcome (PICO) model[34]. As such, the PICO terms for this systematic re- view were as follows:

P: endosomal/intracellular trafficking network. I: HSV1 and HSV2.
C: uninfected cells.
O: alteration of the intracellular/endosomal network.

The “O” term herein refers to modifications in the network’s organellar systems, com- pared to normal, including an increase and/or decrease in the number of organelles, an increase and/or decrease in organellar signalling proteins, an increase and/or decrease in the speed at which organelles move within the cell, a change in the concentration of organelle contents, a difference in the permeability of organelle membrane or an enlarge- ment and/or shrinking of the organelles in size. During search term testing, inclusion of C & O terms was overly limiting such that known reports relevant to the research question were missed. As such, the final searches only used P & I based terms.

The following search terms were used:

> endosome*, trans-golgi network, endocyt*, transport vesicles, endoplasmic reticulum, golgi apparatus, lysosom*, transcytosis, exosom*, exocytos*, pinocytos*, rab gtp-binding proteins, rab, rab* AND herpes simplex, herpesvirus 1 human, herpesvirus 2, human, hsv.

*For full search term descriptions and database listings see the supplementary information*.

### Eligibility Criteria

Search results were assessed for any mention of intracellular trafficking components and HSV-1 and/or HSV-2. Studies were excluded if they were not original primary re- search, were not yet peer-reviewed, unavailable in English or did not assess human neuronal cells (Table 1).

**Table 1.** Search Eligibility Criteria.

|  | Inclusion | Exclusion |
| --- | --- | --- |
| **Population** | Primary or cultured human neuronal cells. Cell lines that can be reasonably considered neuronal due to origin or the presence of neuro-specific cellular components. | Non-neuronal human cells/tissues. Non-human cells/tissues. |
| **Intervention** | Studies specifically assessing HSV-1 and/or HSV-2 infection. | Studies assessing other herpes viruses, or non-herpetic viral infection. |
| **Observations** | Studies presenting primary data on intracellular trafficking components in response to or in the presence of HSV. | Studies presenting data on any other cellular component/system in response to HSV including intercellular trafficking/communication. |
| **Publication type** | Any report presenting original primary research data. | Review articles / experimental proposals / editorials / commentaries |
| **Language** | Studies published in English. | Studies not published in English. |
| **Peer review** | Studies published in a peer-reviewed journal. | Studies not published in a peer-reviewed journal, including pre-prints. |

The Web of Science, Scopus and Medline databases, were searched on the 5th July 2021. This was repeated on the 28th of December 2021, the 10th of June 2023 and the 18th of January 2023 to assess any newly published reports. Using our broad search terms, 6292 records were initially identified (Fig.2). Following automated duplicate removal by EndNote software, and manual duplicate removal, 4185 reports were screened by title, then by abstract content. 48 articles were screened based on their full text. Additional eligible studies were located by assessing the bibliographies of reports that met final eli- gibility criteria during the systematic search.

**Figure 2.**
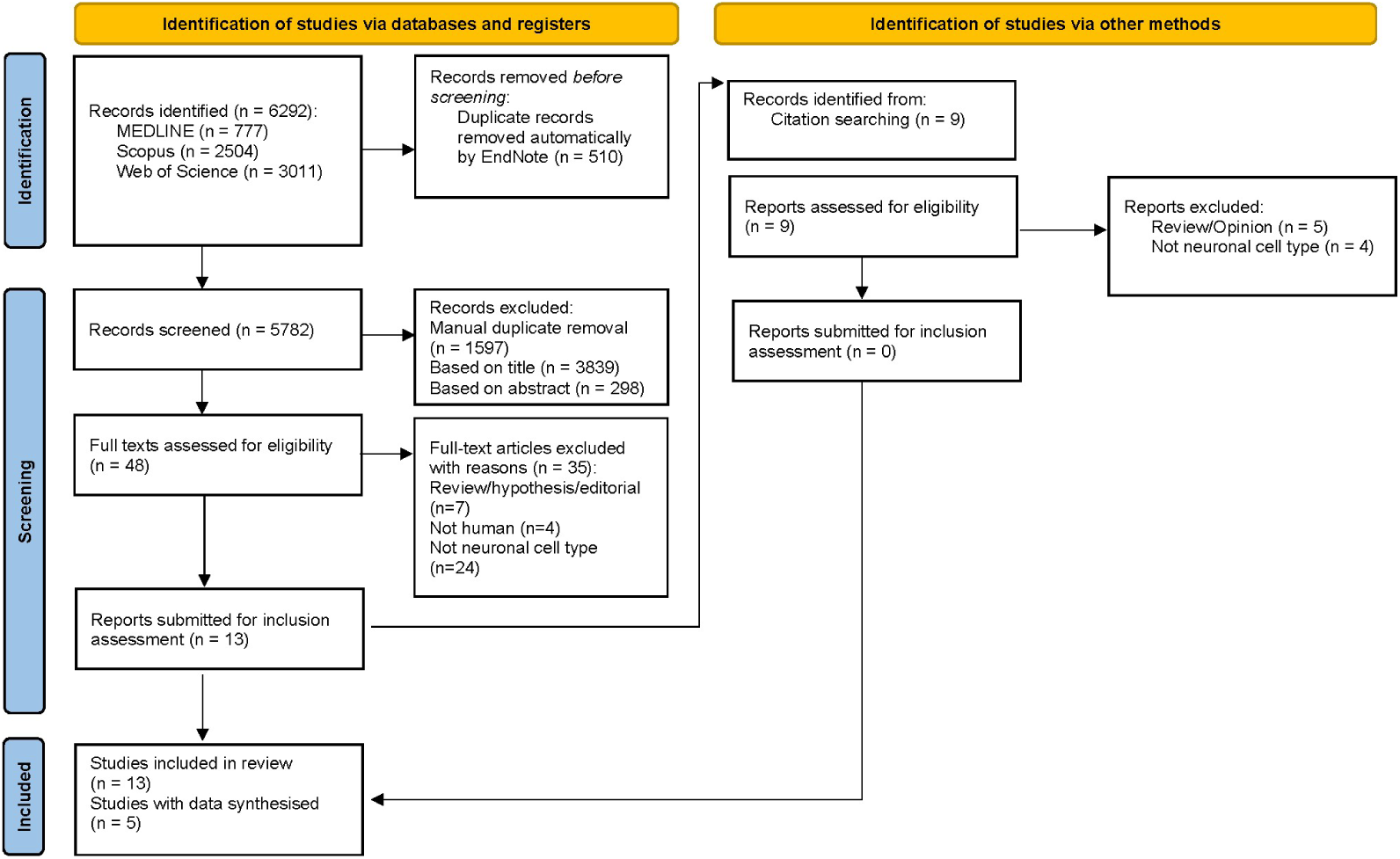
Flow chart of screening, and criteria for inclusion and exclusion of articles

To address the aims of this study, eligible reports were further screened based upon the presentation of observations/data of modification of the HINT system (described above “O search term”) due to HSV1/2 herpes virus infection. Reports that focussed on viral binding sites, transport mechanisms, or the effects of cellular activity on viral particles were excluded from our study.

## Results and Discussion

Our database search produced 13 articles that met the eligibility criteria outlined in Table 2. Of these, only five presented observations/data on effects on HSV on HINT, and will be assessed separately in detail. Six of the remaining eligible reports came from con- temporary work from two separate labs, both of which worked on understanding the mechanisms of HSV transport and assembly within axons [35],[36],[37] and [38],[39],[40]. Movaqar *et al.* (2021) described the effect of autophagic activity on viral proliferation[41], while Musarrat and colleagues (2021) only presented human cell data regarding viral binding proteins[42]. It is notable that in all eligible reports, only HSV-1, not HSV-2 was studied. While these results are interesting in and of themselves, adding to the body of species-specific viral understanding, they do not provide actionable HINT data.

**Table 2.**
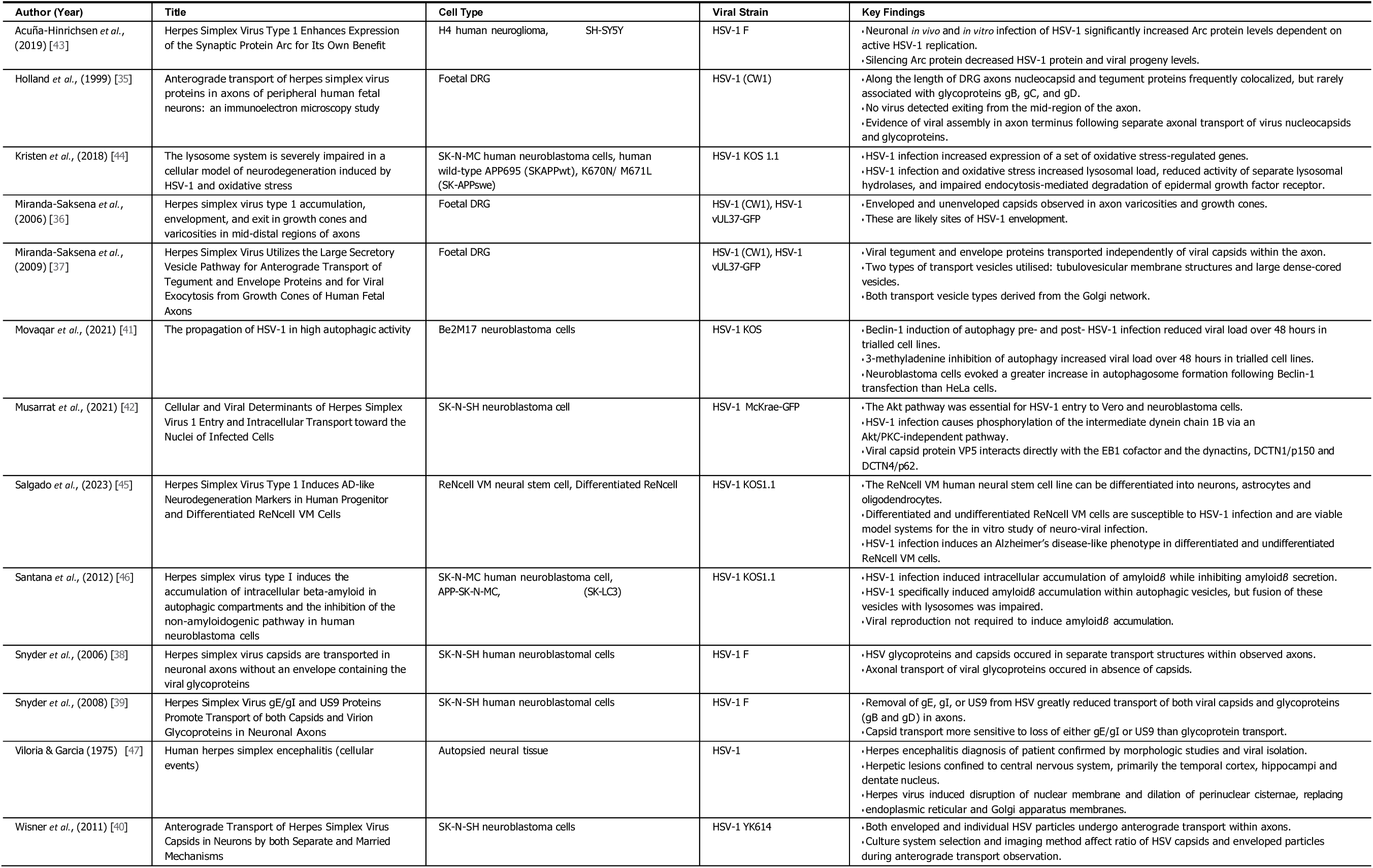
Summary of Studies Meeting Eligibility Criteria.

### HINT Alteration

The five final reports presenting relevant HINT data (see Table 3) consisted of three papers from the group of Aldudo & Bullido which directly investigated the link between HSV and AD[44–46], one independent study investigating the role of HSV-1 in neurode- generation[43], and one clinical report of herpes simplex encephalitis[47].

**Table 3.**
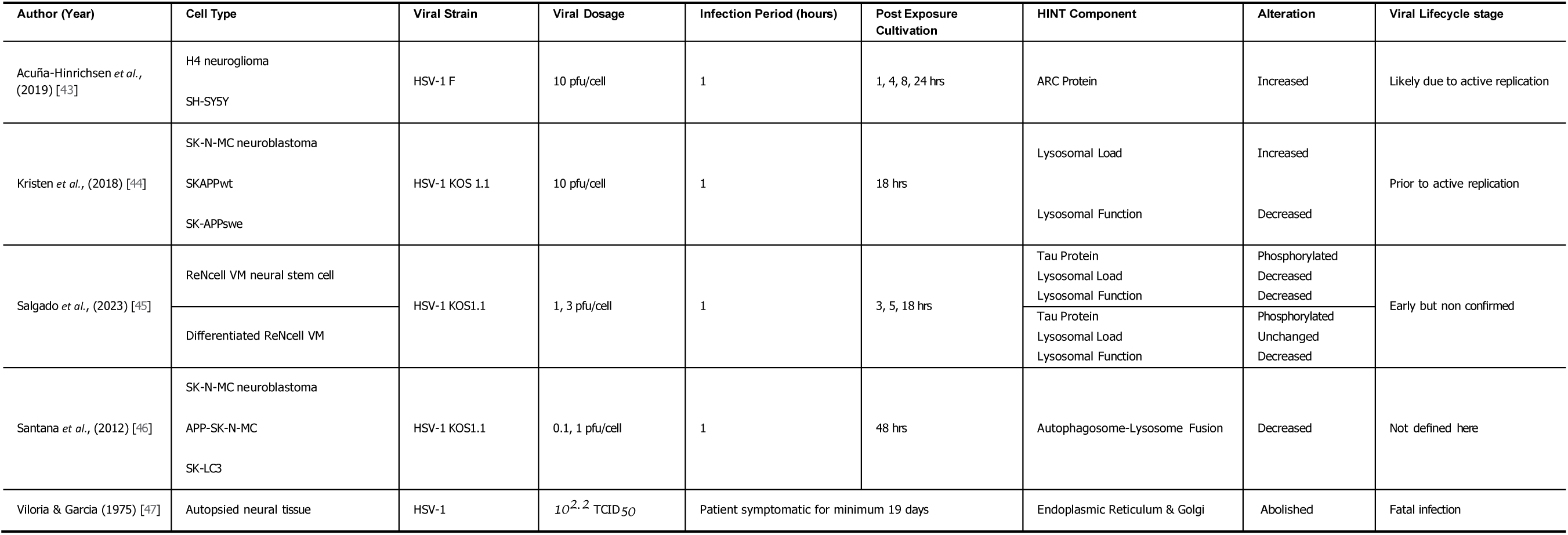
Direct Evidence of HSV alteration of HINT System.

### Lysosome

The collected works of Kristen, Santana and Salgado, coming from the Alduo & Bullido group, all report the effects of HSV-1 on lysosomal health: Specifically, lysosomal func- tion, lysosomal fusion, and lysosomal load.

In 2012 Santana *et al.*[46] reported increased microtubule-associated protein 1 light chain 3 (LC3)-positive autophagic vesicles within neuroblastoma cell lines in response to HSV-1 infection, with A*β* observed to co-localize with these autophagic vesicles following infec- tion. The effects were HSV-1 specific as they were not replicated following neuroblastoma infection with a Sindbis viral construct. Despite the assumed final merging of autophagic vesicles with lysosomes, no co-localization of A*β* nor LC3 was observed. They concluded that the reduced proteolytic cleavage of intracellular A*β* occurs due to reduced matura- tion of autophagic vesicles, and reduced fusion with lysosomes.

Using microarray gene expression analysis on oxidative stress regulated genes in HSV- 1 infected neuroblastoma, Kristen *et al.* (2018)[44] revealed five significantly enriched genes linked to lysosomal system function; Cathepsin F (CTSF), lipase A (LIPA); ATPase H+ transporting accessory protein 1 (ATP6AP1); Niemann-Pick disease, type C1 (NPC1); and lysosomal-associated membrane protein 2 (LAMP2). Both HSV-1 infection and acute oxidative stress produced an increase in lysosomal load (the quantity of contained sub- strates). Lysosomal load increase typically results from either increased intake of sub- strates or diminished degradation within the lysosome. HSV-1 infection decreased lyso- somal cathespin activity, suggesting that the increased load was due to reduced function- ality. Additionally, HSV-1 infection impaired receptor-mediated lysosomal degradation of epidermal growth factor receptor (EGFR). While they concluded that infection induced a great strain on the lysosomal system, they believed that the lysosomal defects reported in their cell model of infection were not directly related to altered lysosomal gene expres- sion. The upregulation of lysosomal genes could be a protective response to counteract impaired lysosomal function.

Finally, Salgado *et al.* (2023)[45], assessed differences in HSV-1 infection in cells of dif- fering differentiation state using their ReNcell VM cell model. Again, HSV-1 infection in- creased the number of LC3-positive autophagic vesicles in both differentiated and pro- genitor ReNcell VM cells, as well as decreasing lysosomal cathespin activity in both cell types. Most interesting however, was the apparent decrease in lysosomal load in pro- genitor cells, in conjunction with the lack of any significant change in the differentiated cells.

Lysosomal dysfunction is proposed to be a key aetiological factor in the pathogenesis of several neurodegenerative diseases such as AD and PD, specifically inregards to impaired clearance of disruptive proteins. The accumulation of autophagic vesicles and impaired degradation pathways observed in HSV-1-infected cells closely mirrors the lysosomal deficits seen in these conditions [48, 49]. For example, cells harbouring AD-related mu- tations in presenilin 2 (PSEN2) expressed increased numbers of LC3-positive autophago- somes, with reduced autophagosome-lysosome fusion [50].

Neural progenitor cells (NPC) and mature neurons respond differently to viral infections due to distinct expression profiles of antiviral genes and innate immune responses [51]. NPCs often exhibit higher levels of autophagic activity compared to differentiated cells in order to facilitate expansive proliferation [52, 53] and the cytoskeletal remodelling that occurs during differentiation [54, 55].

### ER and Golgi

Although the case study of Viloria & Garcia [47] is very obviously an acute infection, it did meet all of the inclusion criteria for our study. The main observation of interest from this paper that we extracted is the complete ablation and replacement of the ER and Golgi apparatus following HSV-1.

Corroborating literature exist describing ER and Golgi remodelling by HSV-1 in other cell models [56]. For example, infected Vero cells showed temporary ER enlargement 12 hours post infection, which returned to normal after a further 4 hours [57]. In the same cells, prior to fragmentation, Golgi membranes significantly enlarged at 9, 12 and 16 hours post-infection [57]. Infected HEp-2 cells showed ER membrane concentration around the nuclear rim [58]. In these same cell lines (but notably not in human 143TK- cells), the Golgi apparatus was fragmented and dispersed throughout the cytosol 8 to 12 hours post-infection [59].

It is commonly understood that the ER and Golgi are involved in HSV viron assembly within the cell, often through the provision of necessary membrane components [60–62]. Disruption of ER proteostasis by physiological/environmental insults leads to an ac- cumulation of misfolded proteins, which is an ER stress response. The highly conserved unfolded protein response (UPR) can detect such ER stress, and attempt to correct it by increasing the expression of genes that regulate ER folding and degradation [63]. HSV-1, as well as many other viruses, have evolved to disable UPR during infection [64]. HSV-1 does so by specifically inhibiting UPR *via* the recruitment of protein phosphatase 1 to dephosphorylate eIF2*α*, thereby blocking protein kinase R-like ER kinase (PERK) stress signal transduction [65, 66]. Impaired UPR leads to unchecked protein synthesis and ex- pansion of the ER membrane to compensate [67, 68].

### ARC Protein

Activity-regulated cytoskeleton-associated protein (ARC) is the only neuron-specific element to appear in these results. Rather than being an active trafficking component, ARC is a hub protein that regulates trafficking components essential for neuronal plas- ticity[69]. Most notably with regard to AD research, ARC binds directly to presenilin 1 (PS-1), the primary component of the *γ*-secretase complex [70, 71] which is responsible for cleaving full length amyloid precursor protein into A*β* fragments [72].

Acuña-Hinrichsen *et al.* (2019)[43] investigated whether recurrent HSV-1 viral reactiva- tions at the neuronal level caused neuronal damage/dysfunction or specifically impacted upon synaptic plasticity. They observed that HSV-1-infected mouse brains, primary mouse neurons, H4 neuroglioma cells and neuronally-differentiated SH-SY5Y cells all showed upregulated ARC mRNA expression. Interestingly, within human cells, ARC protein levels increased during the eighth hour of infection, whereas expression in mouse cells had increased rapidly by hour two. In both species the effect was only transitory.

Within the H4 neuroglioma cells, even though ARC plays a role in *α*-amino-3-hydroxy- 5-methyl-4-isoxazolepropionic acid receptor (AMPAR) internalization under basal condi- tions, during HSV-1 infection, AMPARs are redistributed in an ARC-independent manner. Results from mouse cells demonstrated the requirement for active viral replication to induce a sustained increase in ARC levels. Further, ARC distributed in a similar pattern to the Golgi marker GM130, strongly suggesting that ARC is located at the Golgi apparatus during infections, and that ARC protein plays a crucial role in the production of HSV-1 viral progeny.

Further, with regard to HSV-1 mediated effects on ARC, Bergstöm *et al.,* (2021) observed a reduction of ARC mRNA expression in human iPSC-derived cortical neurons 48 hours post-infection [73]. They specifically discussed the study of Acuña-Hinrichsen *et al.,* (2019)[43], but only the data related to primary mouse cortical neurons, and cited experimental de- sign/ species differences as the possible cause for the discrepancy between their results.

However, following a review of the data from both papers, we suggest that the most likely reason for their incongruent results was time point selection, as Acuña-Hinrichsen *et al.,* (2019) present peak ARC mRNA expression at eight hours post-infection followed by decline, whereas Bergstöm *et al.,* (2021) showed only decreased expression at 48 hours. Therefore, there may have been a post-HSV-1 increase in ARC mRNA expression that was missed, followed by a decline to sub-basal levels.

## Conclusion

Due to the specific nature of the original hypothesis, it is perhaps not too surprising that the majority of final synthesised results (Table 3) focus upon investigating the role of HSV in neurodegeneration, mainly AD. What is surprising is the sheer dearth of re- search investigating the effects of HSV-1 infection on human neuronal tissue as it is well known that HSV activity differs between species, cell type, and modality. Even with the limited number of reports presented here, we observed species- and cell type- specific differences in response to HSV-1 infection, a finding that underscores the critical need for cell type- and species-specific research rather than extrapolating from cell line or an- imal study data.

This systematic review highlights the significant gaps in our understanding of the effects of HSV infection on human neuronal cells, particularly regarding the intracellular traf- ficking system. Despite the extensive history of research on HSV-1, there is insufficient primary evidence detailing its specific impacts on the neural endosomal network and in- tracellular trafficking in human neurons. However, the advent and continuous improve- ment of iPSC technologies provides the opportunity to bridge these knowledge gaps [74]. iPSCs can be differentiated into various neural cell types, allowing for more accurate and relevant *in vitro* models of human neurobiology for use in future viral research.

None of the studies discussed herein adequately contributed to the exploration of chronic viral infection and repeated viral reactivation or their roles in neurodegeneration. ARC level increases were likely due to initial active replication, lysosomal/autophagosomal dis- ruptions occurred in early infection or initially prior to active replication, and ER and Golgi disruption was only directly observed in a fatal case of encephalitis. If the hypothesis that long term sub-clinical, non-cytotoxic reactivation of HSV can instigate neurodegenerative disease processes is to be maintained and further posited, new specifically designed ex- periments must be conducted.

While current evidence points to significant effects of HSV-1 on neuronal intracellular traf- ficking systems, primary evidence is still lacking in detail and scope. Findings from our review advocate for increased utilization of cell- and species-specific models to establish more stable foundations for hypotheses regarding HSV involvement in neurodegenera- tion and brain ageing.

### Limitations

This systematic review has several limitations that should be considered when inter- preting the findings. Firstly, the limitation of investigating only studies written in English may have constrained the total number of studies assessed, and introduced a potential bias by excluding relevant research published in other languages, leading to an incom- plete representation of the existing research. Secondly, the heterogeneity of studies in- cluded in this review posed challenges for the final synthesis of results and hindered a robust comparison of study quality. This possible variability in experimental rigor could have influenced the overall findings and the strength of the conclusions drawn.

The most notable limitation of this study is the narrow scope of the findings, due to the limited number of eligible studies. Although the initial research question was highly spe- cific, generating truly comprehensive search terms proved challenging given the broad range of components that can be considered part of the HINT system. For instance, a pub- lication that was later found to have relevant cell data [73] was not discovered during the systematic search, likely because the components were reported in the context of ’neu- ronal plasticity’ rather than ’intracellular transport’. However, one must note, while the final number of studies presented was small, a large number of studies were screened but ultimately deemed ineligible.

Lastly, no standard risk of assessment could be conducted due to the small number of final results, the lack of applicable metrics within those results, and the heterogenous mix of study designs.

## Supporting information

PRISMA checklist

## Data Availability

All data produced in the present work are contained in the manuscript

## Acknowledgment

This preprint was created using the LaPreprint template (https://github.com/roaldarbol/lapreprint) by Mikkel Roald-Arbøl.

## Author contributions

Conceptualization: M.B.V.; Methodology: M.B.V, D.B.; Investigation: M.B.V., D.B.; Writing - original draft: M.B.V; Writing - review & editing: M.B.V, M.G.R; Supervision: M.G.R.

## Supplementary

For full database search terms, see appendix 1 below.

## Appendix 1

### Search Strategy

#### Search Strategy Web of Science

TS=(endosom* OR trans-golgi network OR endocyt* OR transport vesicl* OR endo- plasmic reticulum OR golgi OR lysosom* OR transcytosis OR exosom* OR exocy- tos* OR pinocytos* OR rab gtp-binding OR rab OR rab*) AND TS=(herpes simplex OR herpesvirus, human OR hsv*)

#### Scopus

(TITLE ("endosom*") OR ABS ("endosom*") OR TITLE ("trans-golgi network") OR ABS ("trans-golgi network") OR TITLE ("endocyt*") OR ABS ("endocyt*") OR TITLE ("transport vesicles") OR ABS ("transport vesicles") OR TITLE ("endoplasmic retic- ulum") OR ABS ("endoplasmic reticulum") OR TITLE ("golgi apparatus") OR ABS ("golgi apparatus") OR TITLE ("lysosom*") OR ABS ("lysosom*") OR TITLE ("tran- scytosis") OR ABS ("transcytosis") OR TITLE ("exosom*") OR ABS ("exosom*") OR TITLE ("exocytos*") OR ABS ("exocytos*") OR TITLE ("pinocytos*") OR ABS ("pinocytos*") OR TITLE ("rab gtp-binding proteins") OR ABS ("rab gtp-binding proteins") OR TITLE ("rab") OR ABS ("rab") OR TITLE ("rab*") OR ABS ("rab*") AND TITLE ("herpes simplex") OR ABS ("herpes simplex") OR TITLE ("herpesvirus 1, human") OR ABS ("herpesvirus 1, human") OR TITLE ("herpesvirus 2, human") OR ABS ("herpesvirus 2, human") OR TITLE ("hsv*") OR ABS ("hsv*"))

#### Medline

((endosom*[Title/Abstract]) OR (trans-golgi network[Title/Abstract]) OR (endocyt* [Title/Abstract]) OR (transport vesicl*[Title/Abstract]) OR (endoplasmic reticulum [Title/Abstract]) OR (golgi[Title/Abstract]) OR (lysosom*[Title/Abstract]) OR (tran- scytosis[Title/Abstract]) OR (exosom*[Title/Abstract]) OR (exocytos*[Title/Abstract]) OR (pinocytos* [Title/Abstract]) OR (rab gtp-binding[Title/Abstract]) OR (rab [Title/Ab- stract]) OR (rab* [Title/Abstract])) AND ((herpes simplex[Title/Abstract]) OR (her- pesvirus, human[Title/Abstract]) OR (herpesvirus, human[Title/Abstract]) OR (hsv* [Title/Abstract])) Initial search conducted 05-07-2021. Updated searches conducted on 28-12-2021, 10-06-2023, and 18-01-2024.

